# PERCOVID: A Model to Describe COVID Percolation on a Network of Social Relationships

**DOI:** 10.1101/2021.08.31.21262909

**Authors:** Jean-François Mathiot, Laurent Gerbaud, Vincent Breton

## Abstract

We develop a site-bond percolation model, called *PERCOVID*, in order to describe the time evolution of *COVID* epidemics and more generally all epidemics propagating through respiratory tract in human populations. This model is based on a network of social relationships representing interconnected households experiencing governmental non-pharmaceutical interventions. The model successfully accounts for the *COVID-19* epidemiological data in metropolitan France from December 2019 up to July 2021. Our model shows the impact of lockdowns and curfews, as well as the influence of the progressive vaccination campaign in order to keep *COVID-19* pandemic under the percolation threshold. We illustrate the role played by the social interactions by comparing a typical scenario for the epidemic evolution in France, Germany and Italy during the first wave from January to May 2020. We investigate finally the role played by the α and δ variants in the evolution of the epidemic in France till autumn 2021, paying particular attention to the essential role played by the vaccination. Our model predicts that the rise of the epidemic observed in July 2021 will not result in a fourth major epidemic wave in France.

## Introduction

Before the development of an approved vaccine for the prevention of *SARS-CoV-2* infection, and in the absence of curative pharmaceutical options for the treatment of the *COVID-19* pandemic, *Non Pharmaceutical Interventions (NPIs)* have been the only available policy levels to reduce the virus transmission ^1^. In this respect, mathematical modelling has proven to be an important tool in order to guide policy makers for taking the appropriate decisions regarding these *NPIs*. Reciprocally, the wealth of epidemiological data made publicly available since the beginning of the pandemic provides a unique opportunity to understand the main characteristics of the existing models on a deeper ground.

The modelisation of epidemics propagating through respiratory tract in human populations relies on two observations: they propagate through the formation of clusters of increasing size and this propagation is mainly governed by the density and the intensity of the social relationships in the population under study. In the case of the *COVID-19* pandemic, there is a slow continuous transmission combining day-to-day interactions, with highest proportion of asymptomatic cases, and accelerations characterized by the links between super spreader events, combining a larger number of human interactions with high viral emission ^2,3,4^. Although this has been widely recognized during the *COVID-19* pandemic, these two observations are however not properly taken care of in most of *COVID* epidemic simulations.

The *SEIR* compartmental type models ^5^ rely on a global approach with no information whatsoever on the formation of clusters. They only rely on the total density of susceptible, infected or recovered persons over the whole territory under study, independently of their social surrounding. The role of social relationships is embedded in a very general parameter - the so-called reproduction number *R*_*0*_ - which incorporates also epidemiological information like the intrinsic infectiosity and infectious period duration. The characteristic length scale of these models is thus not small enough to be able to follow the epidemic on the long term when the local environment of each person starts to be most important and governs the virus propagation^6,7,8^.

The *Multi-Agent System* models are very detailed simulations of a restricted territory – such as a town - with a rather large granularity - like a building - and a small time scale - like an hour. They require evaluating multiple scenarii and therefore large computing capacities^9^. It is therefore very difficult to extract any generic behavior at the level of a large territory and to identify collective effects like abrupt transitions. The characteristic length scale of such models is thus much too small to follow the epidemic over a long period of time.

These remarks motivated our choice to consider a realistic model at a mesoscopic scale, based on percolation theory ^10^. The formation of clusters is the way percolation models operate, while collective behaviors lead to a sharp transition, the so-called percolation transition. This transition occurs from a *non-percolation* regime with many small clusters to a *percolation* regime with a large cluster at a macroscopic scale, *i*.*e*. with a typical size of 50% of the population or larger. Compared to previous attempts at modeling the *COVID-19* pandemic using percolation-type models ^11^, we advocate in this study its formulation on a *network of social relationships*, emphasizing the role of these relationships in the propagation of the epidemics rather than the geographical proximity of persons. The interpretation of percolation theory in terms of social relationships was already applied to other contexts ^12^. The characteristic mesoscopic scale of such model enables also to follow the epidemic over a very long period of time and the *COVID-19* pandemic provides a tremendous testing ground of its validity over more than 600 days.

We describe in this study the general properties of the *PERCOVID* percolation model, detail its construction and show how the virus propagation displays a sharp transition between a percolation and non-percolation regime depending on the density and intensity of social relationships. The results of the model since the beginning of the *COVID-19* pandemic in metropolitan France are presented: the raise of the first wave in March 2020, the impact of the first lockdown, the second wave during the fall 2020 followed by the second lockdown, the third wave in spring 2021, as well as the influence of the curfews and of *Super-Spreading Events* (*SSE*) in this evolution. We illustrate the role of social relationships by comparing the situation in France, Germany and Italy in a typical scenario for the beginning of the epidemic. We also detail the role played by a progressive vaccination campaign in 2021 together with the influence of the propagation of the α variant. From the whole history of the *COVID-19* pandemic, we can finally predict how the δ variant should propagate in France till autumn 2021. A *Methods* section provides all necessary additional details on the *PERCOVID* model.

## Construction of *PERCOVID*

We constructed *PERCOVID* following the main characteristics of an epidemic propagating through respiratory tract in human populations. It is defined on a lattice in *D* dimensions with, as usual in site-bond percolation theory, sites connected by links. The only geographical proximity which is relevant to the propagation of the virus corresponds to persons sharing the same household. Each site of the network is therefore identified with one, two, or three and more persons belonging to the same household, with a distribution according to the territory under study. The connexions between the sites are represented by the bonds between them. They correspond to the daily social interactions between any person in one household with any person in the neighboring household on the lattice.

We need to emphasize here that a neighbor on our (social) lattice is not necessarily a geographical neighbor: indeed, two persons can have close social interactions while living in geographically distant locations. These social relationships do control the propagation of the epidemic and are embedded, by construction, in *PERCOVID*. They correspond for example to interactions in the professional and school circles, during extraprofessional activities with friends or with the family living outside the household. All these activities should not be considered however on the same footing, as far as the propagation of the epidemic is concerned. We therefore consider a first circle of so-called *essential* activities corresponding to the closest neighbors on the lattice, while *less-essential* activities are associated to a second circle represented by next-to-closest neighbors.

The richness of these daily social relationships depends on the dimension *D* of the lattice. For *D=1* for instance, this corresponds to very limited relationships with only two neighbors. For *D=2*, we start having a second circle of less essential relationships, with four next-to-closest neighbors, and four closest neighbors for the first circle. In order to account for the observed social behaviors in different European countries, we consider in this study a three-dimensional lattice. This implies a maximum of six possible daily interactions in the first circle which may lead to a transmission of a given respiratory virus, and twelve more for the second circle. The situation with *D=4* may correspond to exceptional situations like *Super Spreading Events* for instance, with very rich social relationships within a specific group of persons over a limited period of time.

Each site of the lattice is occupied with a probability *p*. Since the links over the lattice are associated to daily social relationships, this probability is therefore also identified with the mean density of these relationships on the territory under study, normalized to one for a maximum of *18* interactions in both the first and second circles. The value for *p* is adjusted in our model to reproduce the average number of daily social relationships in France and various European countries ^13,14^. Following the classification already used in the *SEIR* epidemic models, any person belonging to a household can be either Susceptible, Exposed, Infected or Recovered. Once infected, a person is either pre-symptomatic, symptomatic or asymptomatic. In the case of *SARS-CoV2*, it can also be vaccinated since January 2021 in France. The model can also take into account the concurrent propagation in the population of up to nine variants.

The time evolution of the epidemic is governed by the probability for an infected person to infect a susceptible one in a daily social interaction. This interaction can mainly occur either with one member of the same household or with someone in its first or second circle of neighbors. This probability depends on two very different parameters: the first one, of epidemiological nature, is associated to the intrinsic properties of the virus and to the viral load of the infected person. It corresponds to the ability of the virus to infect one susceptible person during a social contact, independently of any other considerations. We call it *infectiousness* and denote it by *r*, with values ranging from 0 to 1. The second parameter, of social nature, is related to the *intensity* of the social relationships in the territory under study and is denoted by *q*. It is independent of the *density* of these social interactions represented by *p*. It is related, on the one hand, to the social behavior of the population and, on the other hand, to the social distancing measures taken by the authorities and adopted by the population for reducing the propagation of the virus. A so-called temperature effect is introduced to reduce both the infectiousness of the virus and the intensity of social interactions during summer as life style evolves to more outdoor activities less favorable to virus transmission ^15^. In order to account for exceptional interactions outside the first or second circle, the model also opens the possibility for any person to interact episodically with any person anywhere in the territory under study.

Once a susceptible person is infected with the probability *q*.*r*, an internal clock is started to follow the disease evolution. This evolution is governed by the infectious time *τ*_*I*_ when the infected person starts to be contagious, and the recovered time *τ*_*r*_ when it recovers (or dies). At each iteration step, the status of each member of each household is evolved according to the previous principles, as detailed in the Methods section.

## Results

### Phase diagram

The percolation transition for a given respiratory virus can be easily identified in a phase diagram in the *(p,q)* plane. We define this transition by the critical values of *p* and *q* for which the total number of recovered persons at the end of the epidemic exceeds 50% of the (non-vaccinated) population of the territory under study. *Figure 1* displays the phase diagram corresponding to the *SARS-CoV2* initial strain, for four different vaccination rates. We indicate also on this figure the expected positions corresponding to the social behavior of populations in three European countries (France, Germany and Italy) at the begining of the *COVID-19* pandemic.

**Figure 1:**
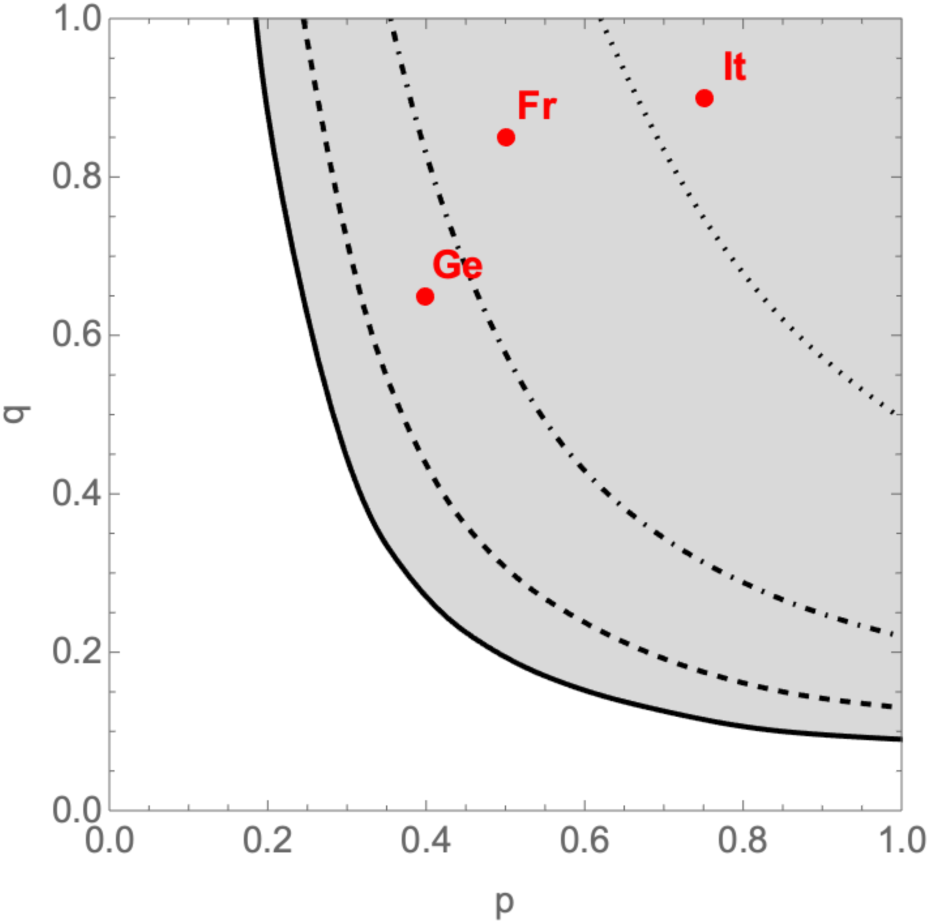
phase diagram of PERCOVID for the SARS-CoV-2 initial strain. The full line shows the limit between the non-percolation and percolation zones, in white and grey respectively in absence of vaccination while the dashed (25% coverage), dot-dashed (50% coverage) and dotted (75% coverage) curves show how the vaccination reduces the percolation zone. The points labeled “Fr”, “Ge” and “It” correspond to the expected position associated to the social behavior in France, Germany and Italy respectively, at the beginning of the epidemic.

According to our model, France, Italy and Germany were all in the percolation zone when the *COVID-19* pandemic started, before any *NPI* was enforced. However, their position reflects the difference in the social behavior of the population in these countries. As we shall see below, this may explain why the impact of the first *COVID-19* pandemic wave was significantly different in these three countries. The relative position of each country with respect to the critical line for a given vaccination rate and a given value of the intensity of social relationships enables us also to predict when the herd immunity should be reached in this country, *i*.*e*. in our study when the non-percolation zone should be reached. The corresponding phase diagram associated to the δ variant is indicated on *Supplementary Figure S1*. Note that the conditions to reach the herd immunity do not depend only on the epidemiological parameters like the infectiousness of the virus or the contamination duration of an infected person, but also on the social behavior of the population through the value of the density and intensity of their social relationships. As a consequence, the rate of vaccination needed to achieve herd immunity does depend explicitly on *q*.

*Figure 2* illustrates the sharpness of the percolation transition, *i.e*. how varying *q* around its critical value, for a given value of *p*, can have a huge impact on the dynamics of the epidemic. While the density *p* of the social tissue is a cultural variable independent of any governmental intervention, wearing a mask and refraining from physical contact translates, in our model, into reducing *q*, the intensity of social interactions. This figure also shows how this transition depends on the vaccination rate. Note that for large values of this rate, the percentage of recovered persons when *q* tends to 1 is not 100% since there remains some islands of isolated sites on the lattice which cannot be reached by the virus.

**Figure 2:**
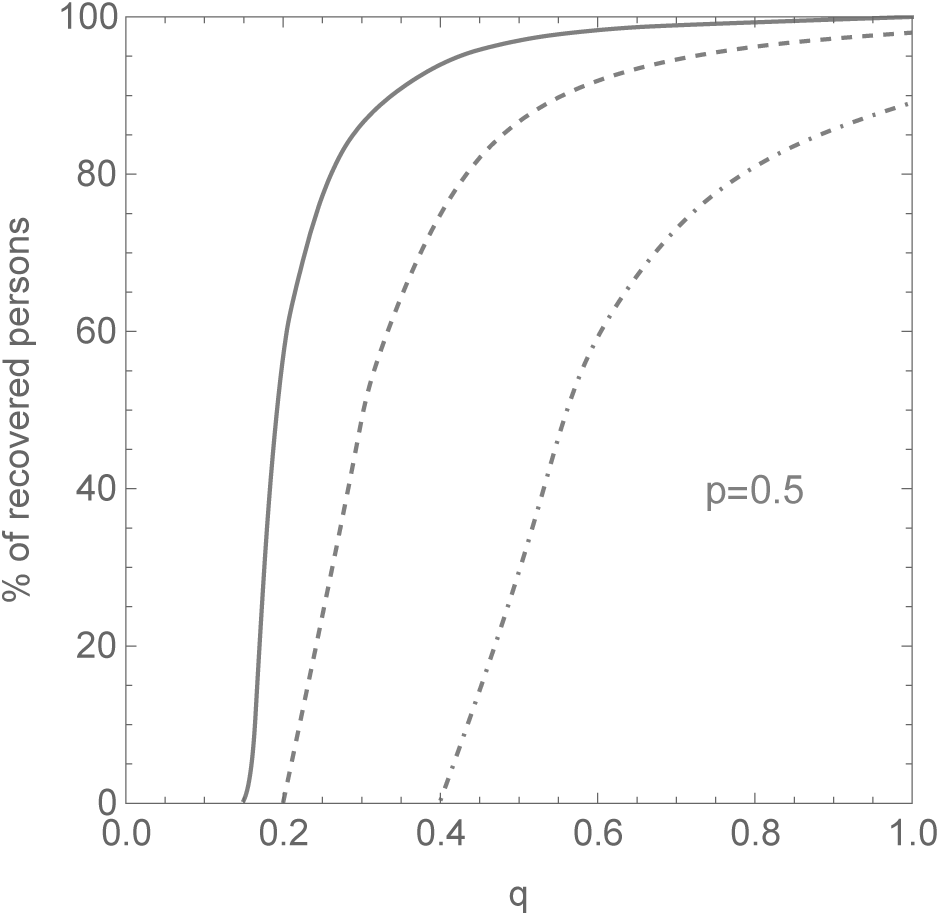
percentage of recovered persons at the end of the epidemic, for the case of metropolitan France (p=0.5), with three different vaccination rates (solid line 0%, dashed line 25% and dot-dashed line 50%) as a function of the intensity q of the social relationships.

### Time evolution of *COVID-19* in France

Since the possible saturation of hospital facilities is the main reason for deciding lockdowns, we choose as major epidemic indicator the number of weekly hospital admissions normalized to 50,000 households. It also better reflects the epidemic evolution than the number of cases that depends on the screening policy. *Figure 3* shows how our model reproduces the normalized weekly hospital admissions due to *COVID-19* pandemic in metropolitan France from December 2019 to July 2021. The simulation is done on a lattice representing the number of metropolitan French households at a scale of *1/10*. It takes into account the influence of the progressive vaccination campaign as well as the propagation of the α and δ variants. *NPIs* endhorsed by the French government are accounted for by changing *q* from its initial value together with possible restrictions to the access to the second circle and to the mobility and contacts ouside the first and second circle, while keeping *p* constant. All details on the choice of the parameters of the simulation are given in the Methods section.

**Figure 3:**
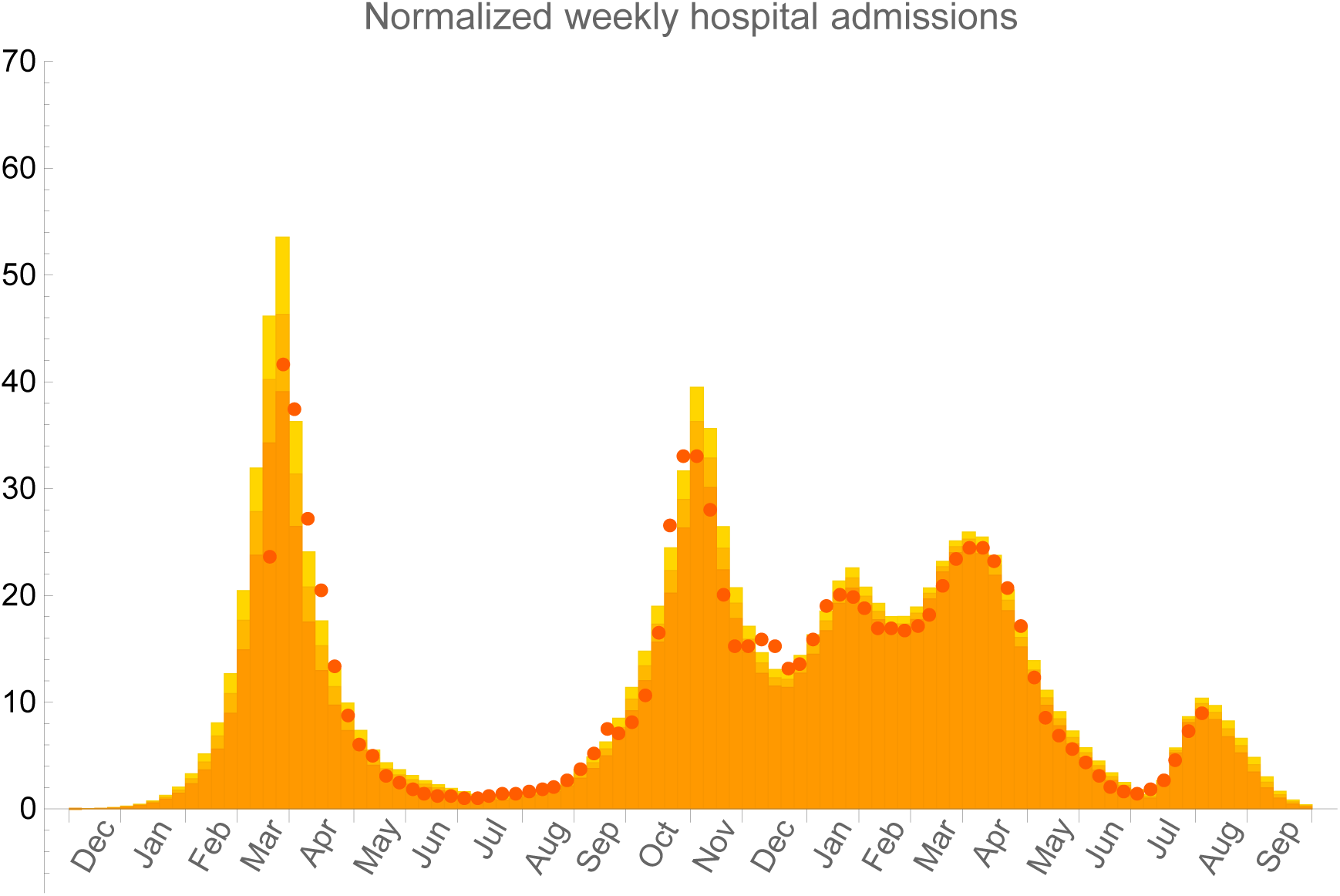
number of weekly hospital admissions normalized to 50,000 households. Orange bars correspond to PERCOVID model prediction averaged over ten simulations and red points to the epidemiological data from Santé Publique France ^18^. Statistical errors at 95% CL are represented by slightly different orange shades.

Our model agrees with the early circulation of *SARS-CoV-2* in France as documented from population-based cohorts ^16,17^. At the scale of metropolitan France, it corresponds to 140 (symptomatics and asymptomatics) infected persons on December 1, 2019. The model confirms the efficiency of the two national lockdowns from March *17*^*th*^ to May *11*^*th*^, *2020* and from October *30*^*th*^ to December *15*^*th*^, *2020* to slow down the virus propagation. The influence of the curfew starting at *6pm* is also seen from the decrease of the hospital admissions in February 2021. The subsequent third wave in March/April 2021 is entirely due to the rapid propagation of the *α* variant, with a larger infectiousness in a context of reduced vaccination coverage.

Although the δ variant will very rapidly represent about 100% of the infected persons due to its large infectiousness, our model predicts only a moderate increase of the number of hospital admissions in metropolitan France in July and August 2021, before it rapidly decreases in September 2021. This absence of a so-called fourth wave in autumn is entirely due to the fact that there are not enough susceptible individuals in the social local environment of any infected one. Due to the natural immunity already acquired by the recovered persons, and the growing vaccination coverage of the population, the virus cannot therefore propagate anymore on a long length and time scale. This corresponds, in terms of the phase diagram discussed above (see *Supplementary Figure S1*), to the non-percolation regime.

### The role of variants in SARS-CoV2 epidemic evolution

Having adjusted our model parameters to reproduce *COVID-19* pandemic time evolution in metropolitan France, we can investigate the role of the various variants in the propagation of the epidemic according to three scenarii:

a. the propagation of the initial virus strain in absence of variants;
b. the propagation of the initial virus strain together with its *α* variant;
c. the propagation of the initial virus strain together with its *α* and *δ* variants.

*Figure 4* shows the number of weekly hospital admissions normalized to 50,000 households for these three scenarii, with three different vaccination strategies. On *Figure 4.a*, we observe a rapid decrease of the hospital admissions starting from February 2021, even in absence of vaccination. The natural immunity of already infected persons, together with a reduced value of the intensity of social relationships through *NPIs* and favorable temperature effect, is however not enough to completely stop the epidemic. It starts to propagate again, although very slowly, at the end of September 2021 with relaxing social distancing steps. The propagation of the *α* variant is responsible for the third wave in April 2021, as shown on *Figure 4.b*. In absence of a vaccination campaign, and because of the increased infectiousness of this variant, the epidemic would have propagated exponentially just at the beginning of summer time, when the intensity of social relationships increased significantly, together with the increase of travels over the whole territory. The effect of a progressive vaccination campaign in March, April, May and June is clearly shown on *Figure4.c*, as compared to *Figure 4.b*. This campaign is however still not enough to completely stop the propagation of the δ variant since it starts to increase exponentially if the vaccination campaign is stopped together with the increase of travels and social exchanges during summer.

**Figure 4:**
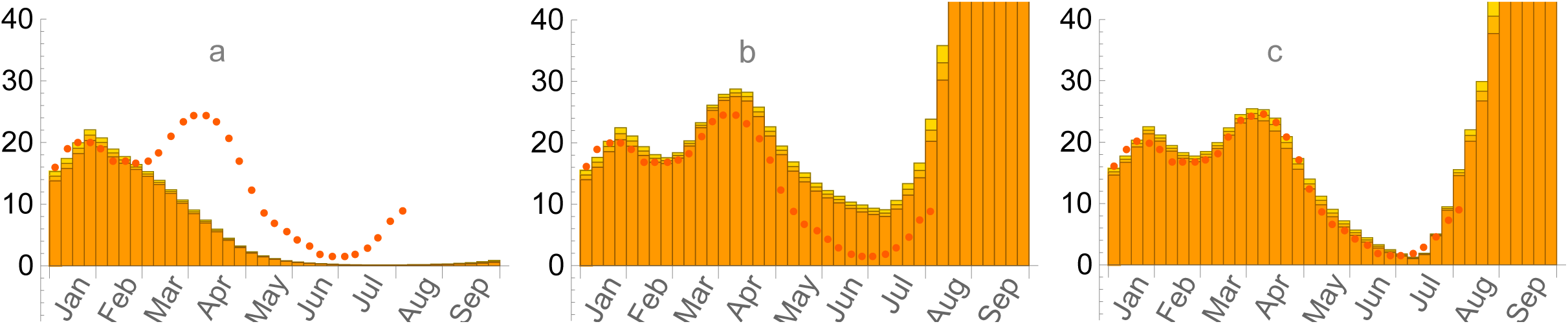
number of weekly hospital admissions normalized to 50,000 households in three typical scenarii; a) with the SARS-CoV-2 initial strain only and no progressive vaccination campaign; b) with the propagation of the α variant and no progressive vaccination campaign; c) with the propagation of the α and δ variants and progressive vaccination campaign up to the beginning of the δ variant propagation only. The red points correspond to epidemiological data from Santé Publique France ^18^ and statistical errors at 95% CL are represented by slightly different orange shades.

These results clearly show the importance of the social behavior of the population in order to be able to predict the influence of a progressive vaccination campaign. When comparing *Figure 4.c* with *Figure 3*, we can see that a continuous vaccination campaign during summer, together with the natural immunity progressively acquired by the infected persons, are enough to completely stop the propagation of the δ variant, eventhough its infectiousness is much higher than that of the *α* variant. Note also that these simulations do not suppose any new *NPIs* in addition to those taken during summer 2020. According to our model, the population fraction either vaccinated or already infected and therefore naturally immunized is around 20% at the beginning of the α variant propagation, and 55% at the beginning of the δ variant propagation.

### Typical expected evolution of the epidemic in France, Germany and Italy

From the phase diagram indicated on *Figure 1*, we can see that the points representing the social behaviors in France, Germany and Italy are all in the percolation zone at the beginning of the epidemic, but in rather different positions. Compared to France, the intensity and density of social relationships is smaller in Germany, and higher in Italy. While the value of the density of social relationships, *p*, can be extracted from literature ^13^, the value of their intensity, *q*, is an educated guess as compared to the value taken for France, with slightly more intensity in Italy and relatively less in Germany, due to different social behaviors. In order to illustrate the influence of these different social behaviors on the propagation of the virus, we consider the evolution of the epidemic till May 2020 with the same confinement restrictions as in France. This scenario highlights the essential role played by the density and intensity of the social relationships in these countries, prior to the subsequent changes of these behaviors following the various *NPIs* enforced by their respective governments. The evolution of the epidemic in Germany, France and Italy, as shown on *Figure 5*, is a direct consequence of the different position of the points associated to these countries in the phase diagram of *Figure 1*. This induces a very slow propagation of the epidemic in Germany, and a much faster one in Italy. Although all points associated to these countries in the phase diagram lie in the percolation zone, the time needed to get an infection propagating on a large scale depends on how far they are from the critical line. Any new enforced *NPIs*, like a confinement period for instance, will thus stop the epidemic at a different stage of its propagation, very soon in Germany and much later in Italy. These patterns correspond qualitatively to what has been observed during the first few months of 2020 in these countries^19^.

**Figure 5:**
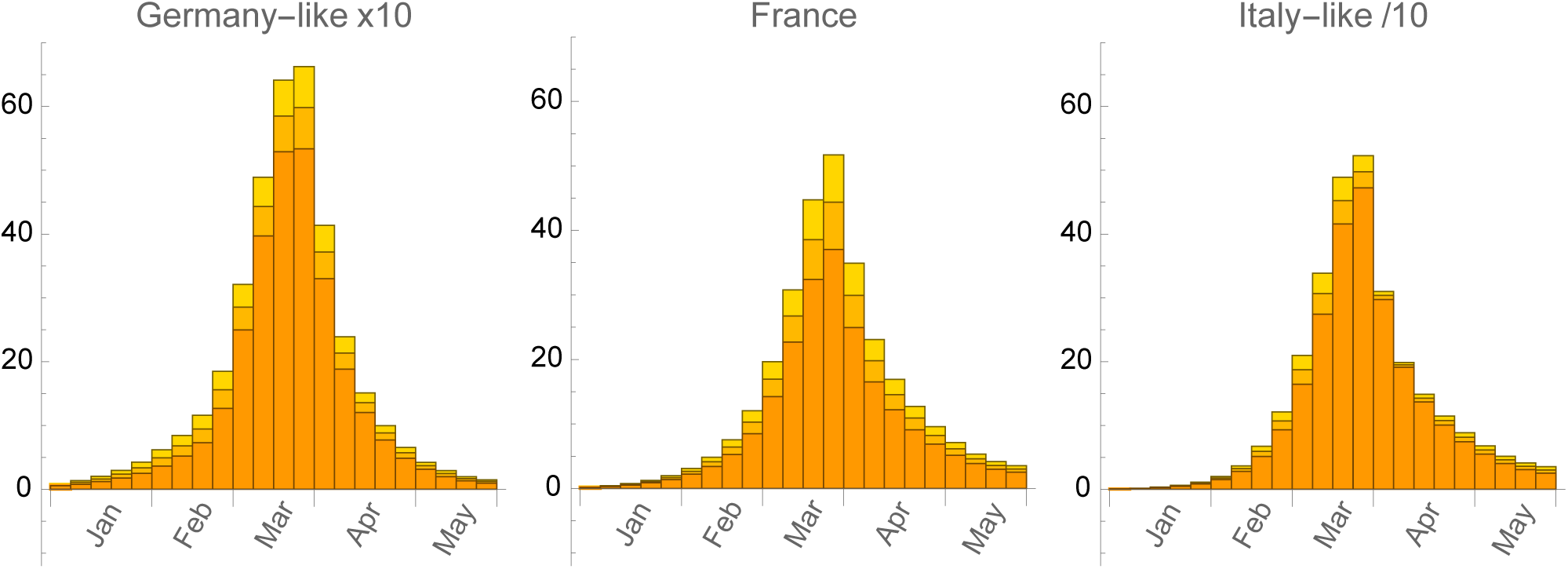
number of weekly hospital admissions normalized to 50,000 households in Germany (multiplied by 10), France and Italy (divided by 10), from left to right respectively, in a typical scenario with no Super Spreading Event, with the same initial conditions, with the same restrictions from the first confinement, and with initial parameters taken from the social behavior in each country prior to any NPIs. Statistical errors at 95% CL are represented by slightly different orange shades.

### The effective reproduction number R

Knowing for each person in each household the whole history of its infection pattern, thanks to the triggering of its internal clock when it gets infected, we can directly calculate the so-called effective reproduction number *R* at any time, as shown on *Figure 6*. This number corresponds to the average over the number of secondary infections an infected person can induce during the period it is contaminant. The basic reproduction number *R*_*0*_, corresponding to the value of *R* at the begining of the epidemic, is not a free parameter in our model, as it is the case in the commonly used *SEIR* models. It depends implicitly on many parameters of quite different nature, like *D, p, q, r* and *τ*_*r*_-*τ*_*i*_.

**Figure 6:**
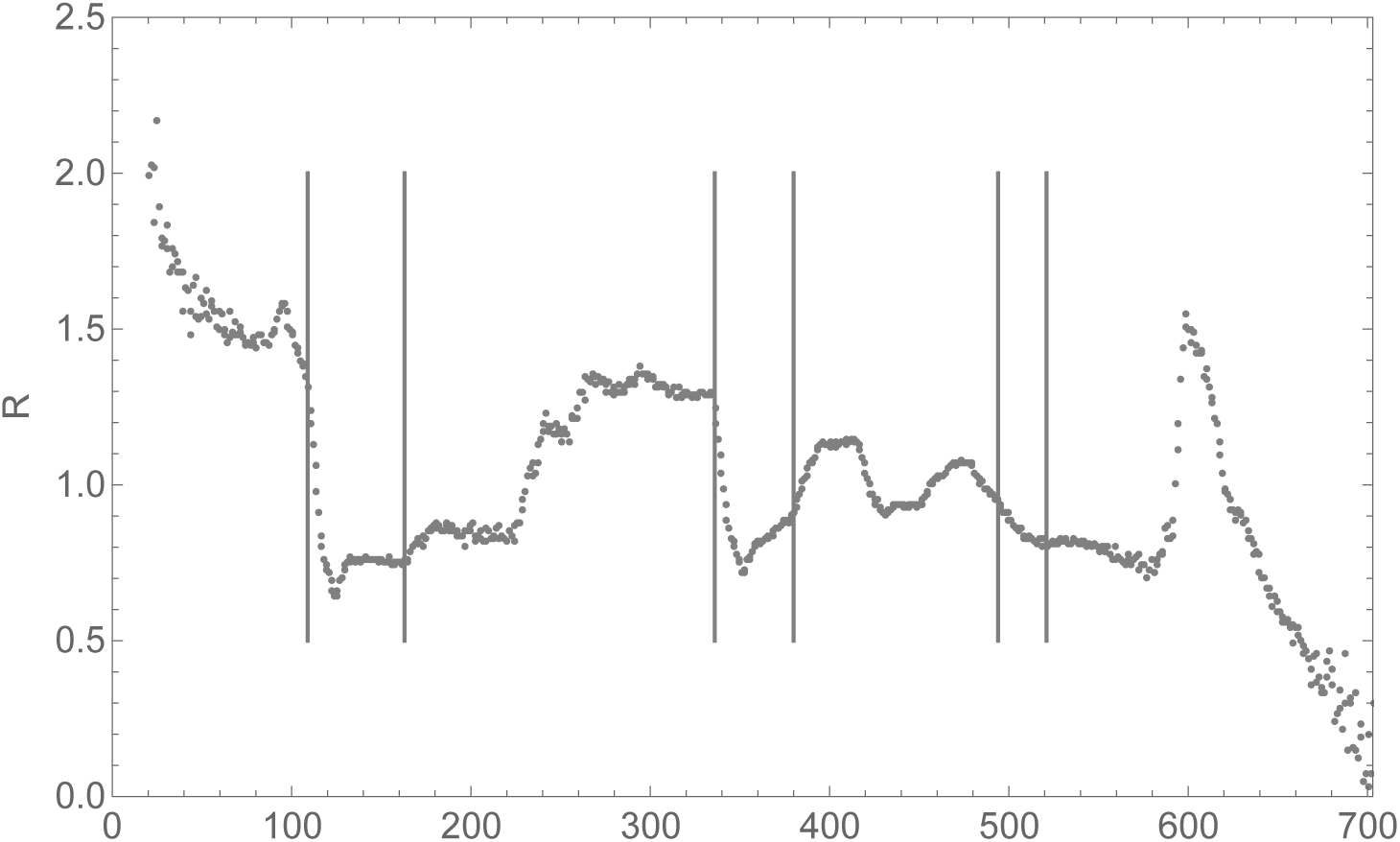
time evolution of the effective reproduction number R during the COVID-19 pandemic in France. The origin of time, t=0, corresponds to December 1,2019. The vertical bars correspond to the beginning and end of the three confinement periods in France.

This figure shows expected features like the rapid decrease of *R* during the beginning of the first and second confinement periods. Note also the peak in *R* just before the first confinement period. It corresponds to the extra infections due to the *SSE* in Mulhouse ^17^. Since the value of *R* was already high at that time, the *SSE* had no large influence on the evolution of the epidemic at country scale. This figure shows also two interesting features as far as the propagation of the variants is concerned. Firstly, the third confinement period had a limited impact for reducing R. It also shows that this observed reduction already started *before* the occurrence of this third confinement period, indicating that the propagation of the α variant was already decreasing in metropolitan France at that time. Secondly, the rapid growth of *R* around day 600 illustrates the very rapid growth of *SARS-CoV-2* infections due to the highly contagious δ variant. However, our model predicts that the progress in the vaccination coverage of the french population, together with the acquired natural immunity will limit very efficiently its impact, leading to a rapid decrease of *R* without the need of any new *NPIs*.

### Perspectives

The *PERCOVID* model provides a simplified but realistic account of the *COVID*-*19* pandemic in metropolitan France. This model emphasizes the importance of the local social environment of each person in a household. This local environment is well accounted for in a site-bond percolation model in terms of the density of social relationships and their intensity. This leads in particular to a sharp percolation transition which may be used as a guiding principle in order to settle appropriate *NPIs* to reduce, and ultimately stop, the epidemic. Our study should be considered as a very first stage of a new paradigm in order to understand epidemics propagating through respiratory tract in human populations, complementing the usual modelisation strategies of epidemics.

From the results presented in this work and the first experience gained in the determination of the parameters associated to the social behavior of the French population, we can make the following statements for the expected situation in autumn 2021. Although any new variant may have an increased infectiousness as compared for instance to the δ variant, it can only spread on a large scale if there is enough space to propagate around each infected person. This should happen if any of the two following conditions is met:

- the variant escapes largely from the protection of the vaccination and of the natural immunity acquired by the already infected persons;
- there is no continuity of the vaccination strategy, if needed from epidemiological studies, like for instance no third dose to extend the period of immunity for the already vaccinated persons or no vaccination for the already infected persons.

Our model is a first attempt in incorporating explicitly the density and intensity of social relationships for the understanding of epidemic propagating through respiratory tract in human populations. These parameters have been adjusted to account for the full evolution of *COVID-19* pandemic from December 2019 to July 2021. They should however be extended in forthcoming studies incorporating for instance a stratification in age in each household in order to be able to make accurate predictions on admissions in Intensive Care Units.

## Data Availability

ALl data referred to in the manuscript are available online.

## Author Contributions

Conceptualization, VB and JFM; methodology, VB, LG and JFM; Code programming, JFM ; writing original draft preparation, VB and JFM ; writing, review and editing, VB, LG and JFM; visualization, JFM; supervision, JFM; project administration, VB, LG and JFM. All authors have read and agreed to the published version of the manuscript.

## Acknowledgements

The authors acknowledge the support of Sorina Camarasu Pop, Jérôme Pansanel and Andrei Tsaregorodtsev for code deployment on France Grilles through SCIGNE platform resources using the VIP portal and the Dirac service.

## Methods

### Parameters of the model

We classify the various parameters of our model into three categories: the epidemiological parameters, the parameters associated to the social behavior of the population in the territory under study and the parameters associated to the mobility of the persons travelling to or within this territory. These parameters are presented in *Supplementary Table S1*. The rate of intra-household infections is determined by the value of *q*_*i*_ and the number *N*_*i*_ of daily intra-household social contacts leading to a potential infection. By construction, we take *q*_*i*_ =*1* as intra-household interactions are the closest. This fixes the general scale for the determination of the intensity of social relationships. The percentage of intra-household infections, as compared to the total number of infections computed in our model, was about 23% just before the first confinement period in metropolitan France, in good agreement with the literature ^20^.

The parameter *p* associated to the density of social relationships is extracted from a population-based contact survey in various European countries^13,14^. For France, the parameters associated to the mobility outside the first and second circle is fixed according to an educated guess, while the intensity of the social relationships is fixed in order to get a consistent description of the *COVID-19* pandemic for more than 20 months. These consistency constraints are strong given that *q* should take a value between *0* and *1* (maximum for intra-household social contact). We assume also that *q* should not be too small (less than *0.20* for instance) in order to keep the possibility to have a very strict limitation of social contacts, comparable to those taken by Chinese authorities in Wuhan at the beginning of the *COVID-19* pandemic. These considerations do fix also the value we should take for the *SARS-CoV2* infectiousness *r*, since the probability for an infected person to contaminate a susceptible one in a daily social interaction is given by *q.r*. To take into account a reduced virulence of the virus during summer season, we introduced a reduction factor *f*_*T*_, with a gaussian time dependence ^15^.

**Supplementary Table S1:** list of all the parameters used to describe COVID-19 pandemic in France. Parameters impacted by governmental NPIs are shown in bold face in the last column. References used to fix epidemiological parameters are given in the last column also. All time scales are given in days.

| *Epidemiological parameters* |  |  |
| --- | --- | --- |
| $(r, r_\alpha, r_\delta)$ | Infectiousness for SARS-CoV2 initial strain and its two variants $\alpha$ and $\delta$ | $(0.14, 0.25, 0.38)^{21,22}$ |
| $dr_{as}$ | Reduction factor of the infectiousness for asymptomatic persons as compared to symptomatic ones | $0.5^{20,23,24}$ |
| $(\tau_i, \sigma_i)$ | Elapsed time from being infected to becoming contagious, with standard deviation | $(3.5, 1.5)^{25}$ |
| $(\tau_r, \sigma_r)$ | Elapsed time from being infected to the end of the contagiousness period, with standard deviation | $(14, 3)^{25}$ |
| $(\tau_r, \sigma_r)^v$ | Elapsed time from being infected to the end of the contagiousness period, with standard deviation, for the $\alpha$ and $\delta$ variants | $(15,3), (16,3)$ |
| $(\tau_h, \sigma_h)$ | Elapsed time between infection and hospital admission, with standard deviation | $(15,3)^{26}$ |
| $(p_h, \sigma_a)$ | Fraction of infected persons requiring hospital admission, with standard deviation | $(3.5\%, 1\%)^{27}$ |
| $(p_h, \sigma_a)^v$ | Fraction of infected persons requiring hospital admission, with standard deviation, for the $\alpha$ and $\delta$ variants | $(5\%, 1\%), (7\%, 1\%)$ |
| $(\tau_s, \sigma_s)$ | Incubation time, with standard deviation | $(5,3)^{28,29}$ |
| $p_s$ | Rate of symptomatic persons among infected persons | $50\%^{23}$ |
| $f_T$ | Reduction factor of infectiousness in mid-summer | $0.5^{15}$ |
| $\sigma_T$ | Standard deviation for the time dependence of infectiousness | 45 |
| *Parameters related to the social behavior* |  |  |
| $p$ | Density of daily social relationships | 0.5 |
| $q$ | Intensity of social relationships | **0.85** |
| $q_i$ | Intensity of intra-household social relationships | 1 |
| $p_c$ | Rate of symptomatic persons staying at home | 50 % |
| $N_i$ | Number of daily intra-household social contacts leading to a potential infection | 5 |
| $dq_{2c}$ | Reduction factor of the intensity of social relationships from the 1<sup>st</sup> to the 2<sup>nd</sup> circle | 0.8 |
| $p_{2c}$ | Probability of having social relationships with the 2<sup>nd</sup> circle | **1** |
| *Mobility parameters* |  |  |
| $N_{ext}$ | Number of daily infections from foreigners visiting metropolitan France | **0** |
| $\rho_{mob}$ | Percentage of persons having social contacts outside the 1<sup>st</sup> and 2<sup>nd</sup> circle | **5 10<sup>-3</sup>** |
| $N_{mob}$ | Number of daily contacts outside the 1<sup>st</sup> and 2<sup>nd</sup> circle, for each person | **5** |

Regarding the vaccination campaign in metropolitan France, we adjust the vaccination rate to the monthly number of persons having full vaccination one week before. Vaccine efficiency, relevant for the ability of a vaccinated person to infect someone else, is set to 90% for both the *SARS-CoV-2* initial strain and its variants.

### Lattice configuration and initial conditions

*Supplementary Table S2* provides details on the configuration of our cubic lattice and the model initial conditions. The initially infected persons from the *SARS-CoV2* strain are randomly generated as symptomatic or asymptomatic with probabilites *p*_*s*_ and (1-*p*_*s*_) respectively. The infection time is also generated randomly within the period [-(τr-τi),0]. In order to account for the expected inhomogeneous distribution of the initially infected persons over the metropolitan French territory in December 2019, we divide our cubic lattice in 3×3×3 sub-regions of equal size. Infected persons are distributed randomly in only five of these 27 regions with a distribution ¼(1,0.9,0.8,0.7,0.6). The number of initial infected persons in our simulation is fixed at 14. For smaller values, large fluctuations in the epidemic time development reduce simulation accuracy. The start of the simulation is adjusted to reproduce the observed peak of hospital admissions during the first wave. Simulating the epidemic evolution in metropolitan France at a scale 1/10 from December 2019 to September 2021 requires about 15 minutes computing time on an Intel Core i3 processor at 1.1GHz clock frequency.

**Supplementary Table S2:**
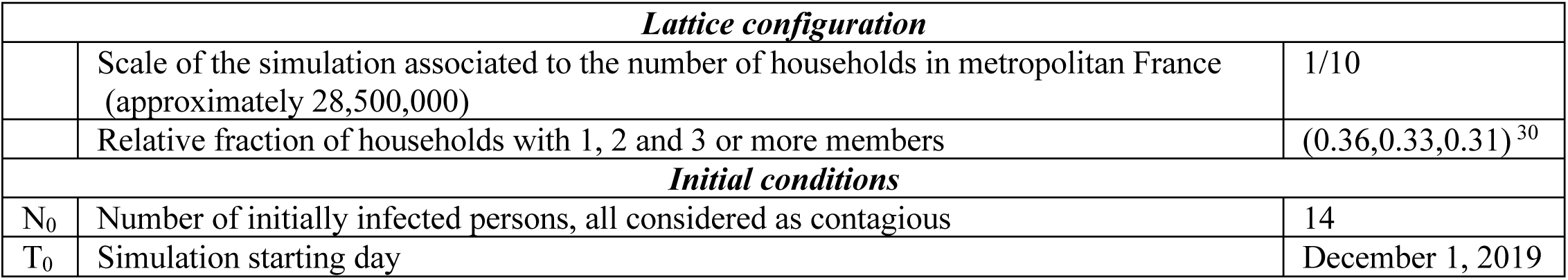
lattice configuration and initial conditions.

As far as the propagation of *SARS-CoV-2* is concerned, and in the absence of a stratification in age that will be considered in a forthcoming study, we assume that a maximum of three persons in each household are susceptible of being infected. This gives a mean number of 1.95 persons per household and corresponds to a population of about 55,500,000 persons susceptible of being infected at the scale of metropolitan France.

The propagation of all *SARS-CoV2* virus, including the initial strain and the α and δvariants, follows the same algorithm. Variant infectiousness has been increased to reflect their higher contagiosity as documented in *Supplementary Table S1*. We consider, for the initial conditions of the variants propagation, the minimum number of daily infections in order to get the variant propagating on a large scale, with a similar inhomogeneous distribution over the metropolitan French territory as compared to the SARS-CoV-2 initial strain. These initial infections are associated to foreigners visiting France. Given the higher infectiosity of the variants, and the mean number of daily relationships we consider in France, the number of these infections for the α and δ variants corresponds approximately to the number of initial infected persons chosen for the *SARS-CoV-2* initial strain. Once the variant propagates significantly in France, additional infections coming from foreigners visiting the country are negligible. The corresponding parameters are indicated in *Supplementary Table S3*.

**Supplementary Table S3:** initial conditions for the introduction of the variants in France.

| *Parameters* | *$\alpha$ variant* | *$\delta$ variant* |
| --- | --- | --- |
| Event start day | February 11, 2021 | July 3, 2021 |
| Event final day | March 11, 2021 | July 24, 2021 |
| *Number of daily infections* | 185 | 290 |

### Time Evolution

Each time step represents one day. This elementary time scale is associated to the definition of the number of mean social contacts per day, as encoded in the parameter *p*. At each time step, we proceed through the following actions:

1. Possible infection in the 1^st^ and 2^nd^ circle as well as intra-household, with symptomatic and asymptomatic rates, for each susceptible person on the lattice.
2. Possible infection outside the 1^st^ and 2^nd^ circle, with symptomatic and asymptomatic rates, for each susceptible person on the lattice.
3. Possible infection all over the country, with symptomatic and asymptomatic rates, from infected foreigners visiting France.
4. Change of status for each person in each household on the lattice according to its internal clock.

### Phase diagram associated to the δ variant

The phase diagram associated to the percolation transition depends explicitely on the infectiousness of the virus. It is indicated, for the *SARS-CoV-2* initial strain, on *Figure 1* while *Supplementary Figure S1* shows the phase diagram associated to the δ variant, corresponding to a very high infectiousness. As expected, the percolation zone extends to a much larger domain in this case, and necessitates a higher reduction of the intensity of the social relationships, and/or a higher vaccination coverage of the population in order to escape from this percolation zone.

**Supplementary Figure S1:**
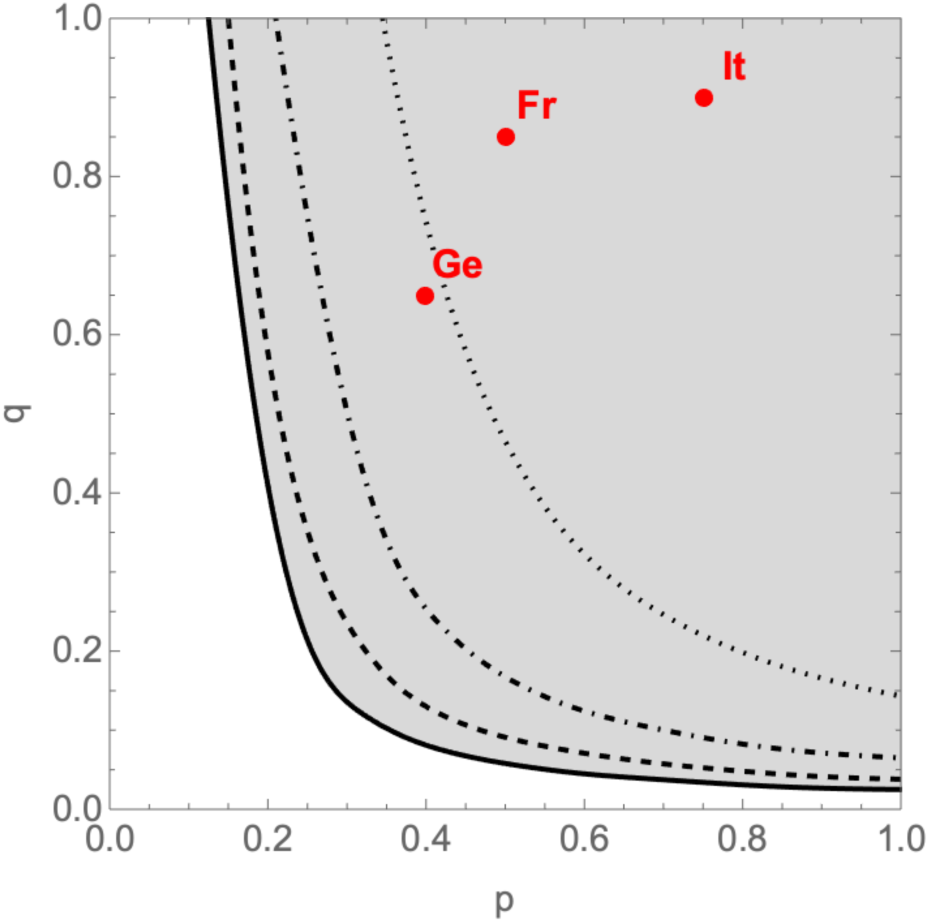
phase diagram of PERCOVID for the SARS-CoV-2 δ variant. The full line shows the limit between the non-percolation and percolation zones, in white and grey respectively, in absence of vaccination while the dashed (25% coverage), dot-dashed (50% coverage) and dotted (75% coverage) curves show how the vaccination reduces the percolation zone. The points labeled “Fr”, “Ge” and “It” correspond to the expected position associated to the social behavior in France, Germany and Italy respectively, at the beginning of the epidemic.

### Changes of social behaviors in France

We detail in this section how the various changes in the social behavior of the French population, due for instance to the governmental *NPIs*, can be accounted for in *PERCOVID* for the period under study. We do not attempt to get a best fit to the data but rather to have a semi-quantitative understanding of the full evolution of the pandemic in metropolitan France. We define in these tables *δρ*_*mob*_ as the ratio of *ρ*_*mob*_ with respect to its value at time *t=0*, as given in *Supplementary Table S1*.

#### Impact of confinement periods

The confinement periods correspond to strong restrictions in the access to the second circle of social relationships as introduced in our model, and in the mobility and contacts outside the first and second circle. These restrictions were very strict for the first confinement period (March–May 2020), and less strict for the second (November 2020) and third confinements (April–May 2021) in metropolitan France. The corresponding expected changes in *δρ*_mob_, *N*_mob_ and *p*_2c_ are documented in *Supplementary Table S4*.

**Supplementary Table S4:** change of social behaviors and mobility parameters during the three confinement periods.

| *Parameters* | *1* | *2* | *3* |
| --- | --- | --- | --- |
| Event start day | March 17, 2020 | October 30, 2020 | April 6, 2021 |
| Event final day | May 10, 2020 | December 12, 2020 | May 2, 2021 |
| $q$ | 0.45 | 0.36 | 0.25 |
| $\delta\rho_{mob}$ | 0 | 0.3 | 0.5 |
| $N_{mob}$ | 0 | 5 | 5 |
| $p_{2c}$ | 0.1 | 0.3 | 0.4 |

#### Epidemic resurgence from the 1^st^ to the 2^nd^ confinement period

The spreading of the *COVID-19* pandemic in France during summer and autumn *2020* is governed by the change of the social behavior of the french population after the first lockdown. This corresponds mainly to an increase of the mobility ouside the first and second circle, as given by the parameters *δρ*_mob_ and *N*_mob_ in *Supplementary Table S5*. The access to the second circle of the social relationships on the other hand is kept the same, and slightly reduced as compared to *1* in order to account for the restriction in the contacts with elderly. In this study, the dates corresponding to the change of social behavior should be understood as indicative of a change of social behavior during summer holidays, the start of the academic year, and the boost of the social as well as economic activities in autumn.

**Supplementary Table S5:** change of social behaviors and mobility parameters between the first two confinement periods.

| *Parameters* | *1* | *2* | *3* | *4* |
| --- | --- | --- | --- | --- |
| Event start day | May 11, 2020 | July 12, 2020 | August 11, 2020 | September 10, 2020 |
| Event final day | July 11, 2020 | August 10, 2020 | September 9, 2020 | October 29, 2020 |
| $q$ | 0.25 | 0.35 | 0.47 | 0.56 |
| $\delta\rho_{mob}$ | 1 | 20 | 15 | 6 |
| $N_{mob}$ | 5 | 10 | 8 | 7 |
| $p_{2c}$ | 0.9 | 0.9 | 0.9 | 0.9 |

#### Curfew periods

Between the second and third confinements, the French government enforced curfews. The curfew periods correspond to slight restrictions in the access to the second circle of the (less-essential) social relationships and in the mobility and contacts outside the first and second circle. This translates in slight reductions in *δρ*_mob_, *N*_mob_ and *p*_2c_, the rate of these reductions depending on the curfew hour (*8pm* for the first curfew and *6pm* for the second), as indicated in *Supplementary Table S6*.

**Supplementary Table S6:** change of social behaviors and mobility parameters during the two curfew periods.

| *Parameters* | *1* | *2* |
| --- | --- | --- |
| Event start day | December 13, 2020 | January 18, 2021 |
| Event final day | January 17, 2021 | April 5, 2021 |
| $q$ | 0.31 | 0.25 |
| $\delta\rho_{mob}$ | 0.9 | 0.6 |
| $N_{mob}$ | 5 | 3 |
| $p_{2c}$ | 0.7 | 0.5 |

#### Epidemic evolution since the 3^rd^ confinement (May 2021)

*Supplementary Table S7* documents the parameters associated to the progressive change in social distancing after the third confinement period, with a smooth transition to parameters similar to the ones taken for the same summer period in 2020. After August 10, 2021, we take similar parameters as the ones in the corresponding period in 2020 (same as column 3 and 4 of *Supplementary Table S5*, except for p_2c_=1.0).

**Supplementary Table S7:** change of social behaviors and mobility parameters after the last confinement period.

| *Parameters* | *1* | *2* | *3* | *4* | *5* |
| --- | --- | --- | --- | --- | --- |
| Event start day | May 3, 2021 | May 19, 2021 | June 9, 2021 | June 30, 2021 | July 21, 2021 |
| Event final day | May 18, 2021 | June 8, 2021 | June 29, 2021 | July 20, 2021 | August 10, 2021 |
| $q$ | 0.25 | 0.25 | 0.25 | 0.30 | 0.35 |
| $\delta\rho_{mob}$ | 0.6 | 0.7 | 0.8 | 10 | 20 |
| $N_{mob}$ | 5 | 5 | 5 | 8 | 10 |
| $p_{2c}$ | 0.5 | 0.6 | 0.7 | 1.0 | 1.0 |

#### Super Spreading Event

A debated issue is whether a *SSE* has played an important role in the emergence of the pandemic in France. The *SSE* which is relevant to our study corresponds to the religious gathering which took place in Mulhouse ^17^ in February *2020*. In this study, we mimic this event by extra contacts from the participants of the meeting, in a restricted sub-region of the lattice corresponding to the size of the city. The parameters of this event are shown in *Supplementary Table S8*. The initial rate of infected participants, as well as the number of extra daily contacts, are deliberately taken as high in order to get an upper limit for the infection.

**Supplementary Table S8:** parameters describing the SSE which occurred in Mulhouse (France) in February 2020.

| *Parameters* |  |
| --- | --- |
| Event start day | February 17, 2020 |
| Event final day | February 22, 2020 |
| Initial infectious rate of the meeting participants | 10 % |
| Total number of participants | 2,000 |
| Total population of Mulhouse city | 110,000 |
| Number of extra daily contacts per participant during the event | 100 |

In view of the early spread of the *COVID-19* pandemic in France ^16,17^, the number of infected participants involved in this event is relatively small at the scale of France. In our simulation for instance, we consider that already 140 persons were infected on December 1, 2019. According to our study, this event had therefore no significant impact on the epidemic nation-wide, but a very significant one on the local spread of the epidemic in Mulhouse and in the east of France during that period.

